# CardioPulmoNet: Modeling Cardiopulmonary Dynamics for Histopathological Diagnosis

**DOI:** 10.64898/2026.02.19.26346620

**Authors:** Tuan D. Pham

## Abstract

**Objective:** This study investigates whether incorporating physiological coupling concepts into neural network design can support stable and interpretable feature learning for histopathological image classification under limited data conditions.

**Methods:** A physiologically inspired architecture, termed CardioPulmoNet, is introduced to model interacting feature streams analogous to pulmonary ventilation and cardiac perfusion. Local and global tissue features are integrated through bidirectional multi-head attention, while a homeostatic regularization term encourages balanced information exchange between streams. The model was evaluated on three histopathological datasets involving oral squamous cell carcinoma, oral submucous fibrosis, and heart failure. In addition to end-to-end training, learned representations were assessed using linear support vector machines to examine feature separability.

**Results:** CardioPulmoNet achieved performance comparable to several pretrained convolutional neural networks across the evaluated datasets. When combined with a linear classifier, improved classification performance and higher area under the receiver operating characteristic curve were observed, suggesting that the learned feature embeddings are well structured for downstream discrimination.

**Conclusion:** These results indicate that physiologically motivated architectural constraints may contribute to stable and discriminative representation learning in computational pathology, particularly when training data are limited. The proposed framework provides a step toward integrating physiological modeling principles into medical image analysis and may support future development of transferable and interpretable learning systems for histopathological diagnosis.

## 1 Introduction

Accurate histopathological assessment is the cornerstone of medical diagnosis and grading, guiding both treatment decisions and patient prognosis [1, 2, 3, 4, 5]. However, manual examination of tissue slides is labor-intensive, subject to inter-observer variability, and often limited by subtle morphological differences among lesions [6, 7, 8]. Recent advances in deep learning have transformed computational pathology by enabling automated feature extraction and classification of histopathological images [9, 10, 11, 12, 13]. Convolutional neural networks (CNNs) have demonstrated high accuracy in detecting malignant patterns [14, 15, 16, 17, 18], while transformer-based architectures have improved contextual reasoning across large-scale tissue regions with clinical records [19], and several areas in histopathological image analysis [20]. Despite these achievements, most current models remain biologically ungrounded, requiring extensive labeled datasets and offering limited interpretability in clinical settings. Moreover, their performance tends to degrade when applied to smaller or heterogeneous datasets typical of cancer cohorts.

To address these challenges, several studies have explored hybrid models combining convolutional backbones with attention mechanisms [21, 22], pretrained CNNs with recurrent neural networks and nonlinear dynamics [23], multi-dimensional feature fusion [24], multi-scale context fusion [25], multi-model feature fusion [26, 27], multimodal fusion [28, 29]. and self-supervised pretraining [30]. While these approaches improve representation quality, they do not capture intrinsic biological processes that naturally balance local and global information flow. The cardiopulmonary system, by contrast, represents an efficient biological mechanism where the heart and lungs operate in dynamic equilibrium through tightly regulated ventilation–perfusion coupling. This interaction ensures stable exchange, adaptability, and resilience under fluctuating physiological conditions—principles highly desirable in neural computation.

Inspired by this mechanism, this work introduces CardioPulmoNet, a physiologically motivated neural architecture that models the interaction between “pulmonary” and “cardiac” subsystems [31] to enhance histopathological image classification. The pulmonary pathway emphasizes local morphological diversity, while the cardiac pathway integrates global contextual coherence. A synchronized multi-head attention mechanism mediates their interaction, emulating cardiopulmonary exchange, and a homeostatic loss enforces equilibrium between the two subsystems. By coupling CardioPulmoNet with a support vector machine classifier, the framework achieves superior performance compared with several pretrained convolutional neural network baselines, particularly for small datasets, demonstrating improved generalization and data efficiency. Furthermore, its dual-state representation offers potential for adaptation as a physiologically informed pretrained foundation model for broader biomedical imaging applications.

This study establishes a physiologically grounded approach to neural network design, bridging the gap between physiological principles and computational intelligence. It provides evidence that modeling dynamic equilibrium, inspired by cardiopulmonary function, can yield stable and generalizable learning behavior for complex histopathological data. The key contributions of this study are summarized as follows.

1. Physiologically Inspired Architecture: A novel deep learning model, CardioPulmoNet, is proposed, inspired by the dynamic coupling between the heart and lungs. The architecture models bidirectional information exchange between two complementary pathways—pulmonary (local) and cardiac (global) to achieve effective and interpretable feature learning.
2. Homeostatic Learning Mechanism: A new homeostatic loss function is introduced to regulate the equilibrium between the dual subsystems, emulating physiological stability in cardiopulmonary interaction. This mechanism enhances discriminative learning while improving robustness under limited data conditions.
3. Integration with Classical Machine Learning: The deep representations extracted from CardioPulmoNet are coupled with support vector machines (SVMs) for final classification. This hybrid approach leverages the feature richness of deep models and the stability of margin-based decision boundaries.
4. Application to Histopathology: The framework is applied to the classification of histopathological images, achieving superior performance compared with multiple pretrained CNN baselines in terms of accuracy, sensitivity, and accuracy.
5. Potential as a Pretrained Foundation Model: The dual-state feature representation learned by CardioPulmoNet exhibits strong transferability, suggesting its potential development as a physiologically informed pretrained model for broader biomedical imaging applications.

The remainder of this paper is organized as follows. Section 2 presents the methodological framework underlying the proposed approach. Section 3 reports the experimental results obtained from three histopathological image datasets. Section 4 discusses the implications of these findings, highlighting their significance and contributions to the field. Finally, Section 5 concludes the paper with a summary of key outcomes and potential directions for future research.

## 2 Methodology

### 2.1 Rationale

CardioPulmoNet is a physiologically grounded architecture that models two interacting subsystems inspired by cardiopulmonary function: a pulmonary (lung) stream that emphasizes local morphological diversity and a cardiac (heart) stream that emphasizes global contextual integration. Information exchange between these streams is implemented via multi-head attention (“alveolar exchange”), while recurrent state updates emulate rhythmic cycles of ventilation and perfusion. A homeostatic loss encourages balanced interaction, and an optional ventilation–perfusion matching term regularizes the ratio of stream activations. Although the proposed framework is applicable to multimodal data, the present work focuses on the image-based case, as demonstrated through experiments on histopathological datasets.

### 2.2 Physiological Analogy

The conceptual foundation of CardioPulmoNet is built upon a physiological analogy between the coupled dynamics of the human cardiopulmonary system and deep neural feature integration. In biological systems, the lungs and heart operate as two interdependent subsystems that exchange oxygen and carbon dioxide to sustain homeostasis. The lungs facilitate ventilation, performing localized gas exchange within alveolar units, while the heart drives perfusion, circulating oxygenated blood throughout the body. Effective function depends on the balance and synchronization of these two processes—a principle known asventilation–perfusion (V/Q) matching.

In CardioPulmoNet, these physiological processes are reinterpreted as a computational metaphor for feature learning in histopathological image analysis. The Lung stream represents local morphological perception, akin to alveolar-level sampling of fine-grained tissue structures. Conversely, the Heart stream captures global contextual coherence, analogous to systemic circulation integrating spatially distributed cues across the tissue microenvironment. The two streams interact cyclically through a bidirectional multi-head attention mechanism that simulates gas exchange, allowing local and global representations to refine one another iteratively.

The homeostatic loss term formalizes the notion of physiological equilibrium: it constrains both streams to maintain stable activation magnitudes around target set-points, mimicking the regulation of oxygen and carbon dioxide levels. Additionally, the ventilation–perfusion loss penalizes deviation from an optimal ratio between Lung and Heart feature activations, ensuring synchronized local–global balance analogous to healthy cardiopulmonary coupling.

Through this biologically grounded design, CardioPulmoNet transcends the purely data-driven paradigm of conventional deep learning by embedding physiological coordination principles into its computational framework. This analogy not only enhances interpretability but also confers robustness and data efficiency, allowing the model to generalize effectively even in limited-data regimes typical of cancer histopathology.

Figure 1 illustrates the conceptual analogy between cardiopulmonary physiology and the CardioPulmoNet architecture, where the interaction between pulmonary ventilation and cardiac perfusion serves as a biological metaphor for the model’s dual-stream feature processing and exchange mechanism.

**Figure 1.**
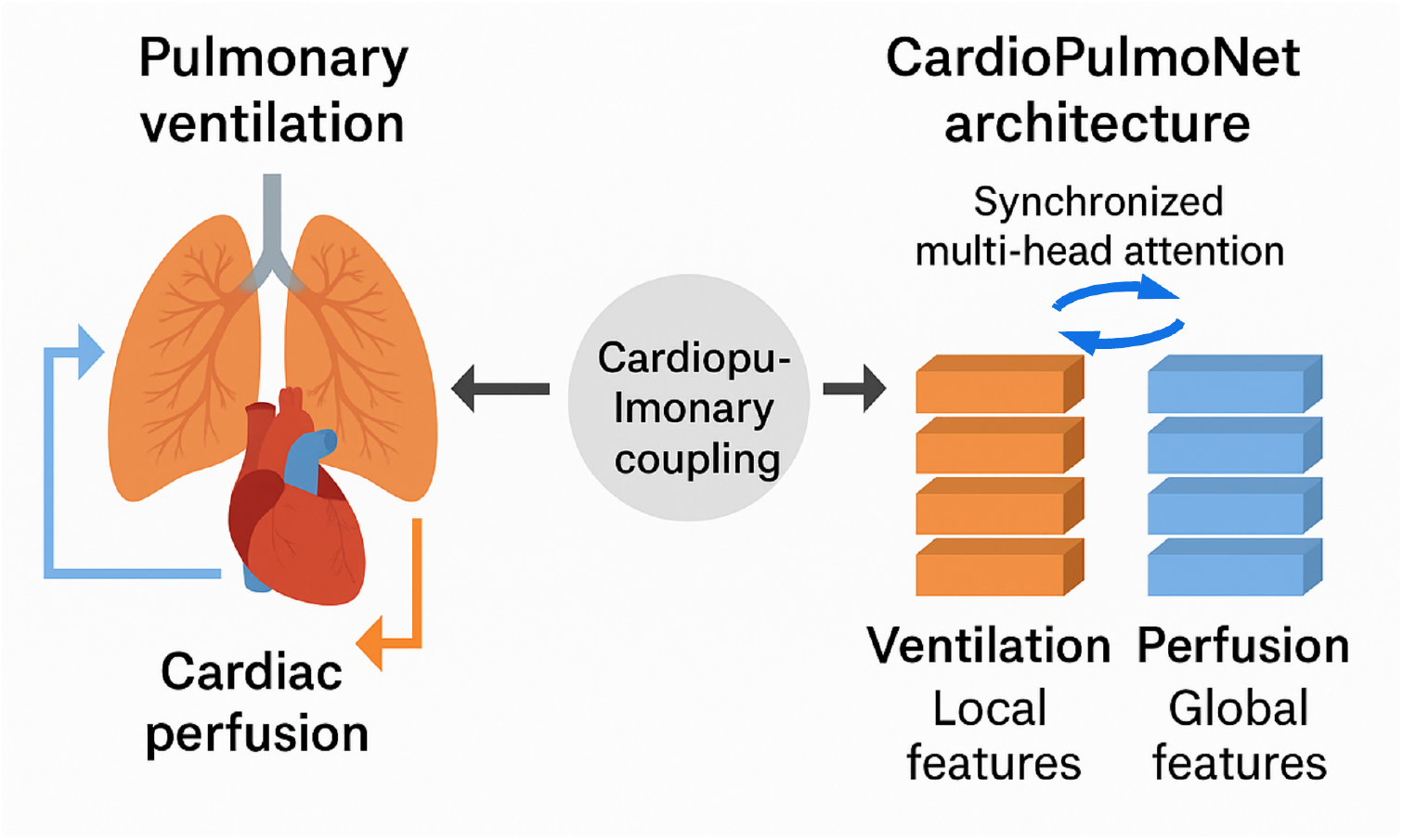
Conceptual analogy between cardiopulmonary physiology and the CardioPul-moNet architecture: The lungs represent ventilation, analogous to local feature encoding in histopathological images, while the heart represents perfusion, corresponding to global feature integration. The bidirectional arrows indicate dynamic interaction and synchronization between these two subsystems, similar to multi-head attention–based information exchange in CardioPulmoNet. This coupling enables balanced and interpretable feature fusion, reflecting the physiological interdependence of pulmonary and cardiac functions.

### 2.3 Notation

Let a minibatch contain *B* images, each resized to *H × W ×* 3. Each image is divided into *S* non-overlapping patches of size *P × P* pixels (*S* = *HW*/*P*^2^). Bold uppercase letters denote tensors, bold lowercase denote vectors, and calligraphic letters denote sets. The main symbols are:

- **X** ∈ ℝ^*B×S×p*^: patch embeddings (*p* is the patch embedding dimension).
- **L**^(*t*)^, **H**^(*t*)^ ∈ ℝ^*B×S×d*^: Lung and Heart states at cardiopulmonary cycle *t* = 1, …, *T*.
- *d*: model width; *h*: number of attention heads; *C*: number of classes.
- [·; ·]: channel-wise concatenation; ⊙: Hadamard product; *σ*(·): logistic sigmoid; softmax(·): row-wise softmax.

### 2.4 Input and Patch Encoding

Images are split into patches and linearly embedded, then projected to the model width:

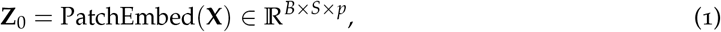

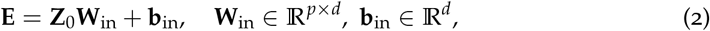

where **W**_in_ and **b**_in_ denote the input projection weights and bias that transform raw patch embeddings into the model’s internal representation space.

The Lung and Heart states are initialized as:

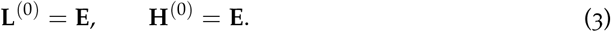

### 2.5 Alveolar Exchange (Multi-Head Attention)

For each cycle *t* = 1, …, *T*, the Lung and Heart streams exchange information using multihead attention (MHA). Given a *query* source **Qsrc** ∈ ℝ^*B×S×d*^ and a *key/value* source **Ksrc** ∈ ℝ^*B×S×d*^, the *i*-th attention head computes:

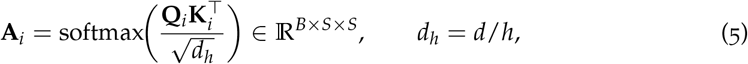

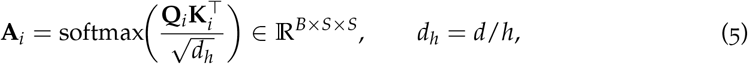

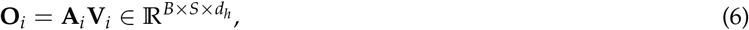

where 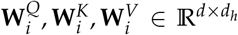 are learnable projection matrices. The *h* heads are concatenated and linearly projected:

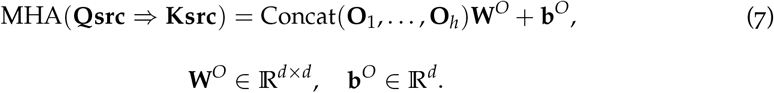

Two bidirectional exchanges per cycle are instantiated as:

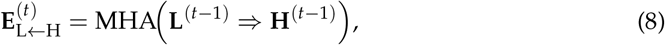

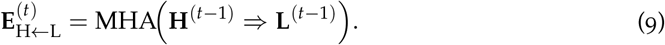

### 2.6 Recurrent State Updates

Each stream updates its state with a gated recurrent unit (GRU) applied token-wise along the spatial dimension *S*. For the Lung stream (input 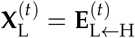, hidden 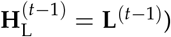):

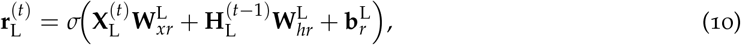

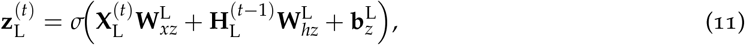

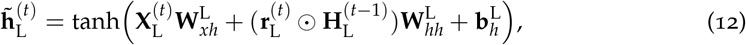

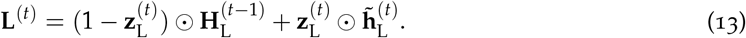

An analogous GRU updates the Heart stream using 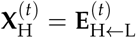 and **H**^(*t*−1)^, with its own parameters {**W**·^H^,**b**·^H^}.

### 2.7 Pooling and Classifier Head

After *T* cycles, the final Heart state is pooled across tokens to form a sample-level descriptor:

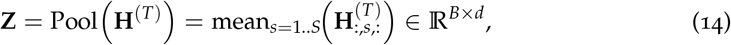

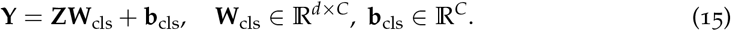

Class probabilities are computed as **P** = softmax(**Y**).

### 2.8 Loss Functions

The total training objective combines a standard classification loss with physiology-inspired regularization terms that encourage stable interaction between the Lung and Heart streams.

#### Classification Loss

Again let **P** = softmax(**Y**) ∈ ℝ^*B×C*^ denote the predicted class probabilities for a batch of *B* samples, where *C* is the number of output classes. Each row **P**_*b*_ = [*P*_*b*,1_, *P*_*b*,2_, …, *P*_*b,C*_] corresponds to the probability distribution over classes for sample *b*, and *y*_*b*_ ∈ {1, …, *C*} denotes the ground-truth class label for that sample. The element 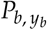 therefore represents the predicted probability assigned to the correct class.

The cross-entropy classification loss is given by

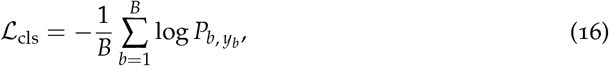

which computes the average negative log-likelihood of the true class probabilities across all samples. This term drives the model to maximize the predicted likelihood for the correct class label.

#### Homeostatic Regularization

To maintain balanced activation magnitudes between the Lung and Heart branches, a homeostatic regularization term penalizes deviations of their mean activations from predefined set-points:

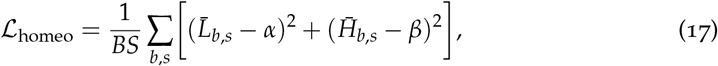

where *α, β* ∈ ℝ are scalar reference levels (set-points) for the mean activations; 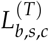 and 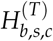 denote the feature activation values at cycle *T*, for batch index *b*, spatial token *s*, and feature channel *c*; 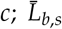 and 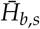 represent the channel-wise mean activations of the Lung and Heart streams for token *s*:

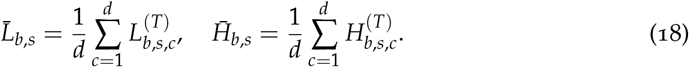

This term enforces a homeostatic balance between the two physiological representations, preventing uncontrolled activation drift in either branch.

#### Ventilation–Perfusion Coupling Regularization

The ventilation–perfusion (V/Q) regularizer enforces a physiological correspondence between the two streams by aligning the ratio of mean Lung and Heart activations:

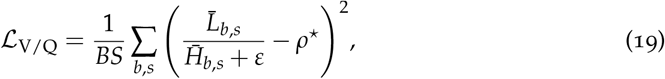

where *ρ*^⋆^ ∈ ℝ^+^ denotes the desired target V/Q ratio, and *ε >* 0 is a small constant to avoid division by zero. This term encourages the dynamic equilibrium between Lung (ventilation) and Heart (perfusion) features, reflecting their physiological coupling.

#### Total Objective

The overall training loss is defined as

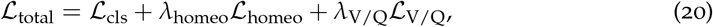

where *λ*_homeo_, *λ*_V/Q_ ≥ 0 are weighting coefficients controlling the contribution of each physiological regularization term. When either regularizer is disabled, its associated *λ* is set to zero.

This composite formulation ensures that CardioPulmoNet not only minimizes classification error but also preserves physiologically interpretable and stable latent representations through homeostatic and coupling constraints.

### 2.9 Training Procedure

Model parameters Θ are optimized using the Adam algorithm with mini-batch training. For each epoch:

1. Sample a batch of images, generate patches, and compute embeddings via Eqs. (1)–(2).
2. For *t* = 1, …, *T*: compute exchanges via Eqs. (8)–(9) and update states using Eqs. (10)–(13).
3. Pool and classify via Eqs. (14)–(15); compute losses via Eqs. (16)–(20).
4. Backpropagate ∇_Θ_ℒ_total_ and update Θ with Adam.

### 2.10 Algorithm

#### Algorithm 1 Forward Pass of **CardioPulmoNet**

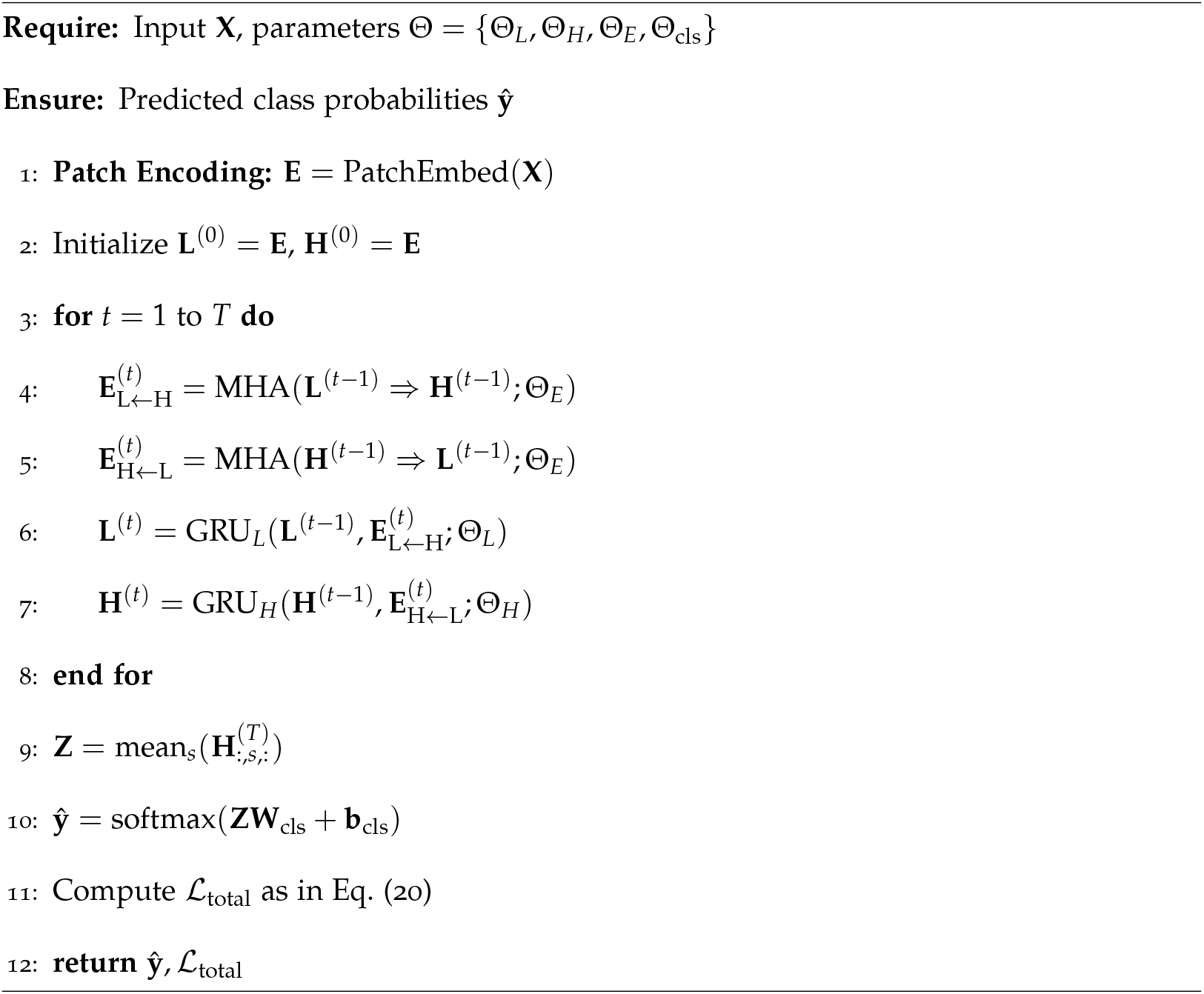

### 2.11 Complexity

With *S*=(*HW*/*P*^2^) tokens and width *d*, each exchange costs *O*(*BS*^2^*d*/*h* + *BSd*^2^), while GRU updates are *O*(*BSd*^2^). The patch size *P* controls the *S*^2^ attention term; *P* is chosen to balance contextual coverage and computational efficiency.

### 2.12 Reproducibility and Pretraining

CardioPulmoNet exposes the pooled descriptor **Z** (Eq. 14) as a reusable representation for classical classifiers (e.g., SVMs) and for transfer learning across histopathology co-horts. This allows its use as a physiologically grounded pretrained encoder by freezing {Θ_*L*_, Θ_*H*_, Θ_*E*_} and fine-tuning Θ_cls_ on new datasets.

## 3 Results

In experiments conducted on three histopathological image datasets, the model configuration was defined as follows. The patch size, model width, number of attention heads, and temporal depth were set to *P* = 16, *d* = 256, *h* = 4, and *T* = 6, respectively. Layer normalization and dropout were applied after each MHA block and before the gated recurrent unit (GRU) inputs to enhance training stability. To enable fair performance comparison, data augmentation was deliberately disabled during training. Training was performed using the Adam optimizer with an initial learning rate of 10^−3^, weight decay of 10^−4^, and minibatch size of 128 for 10 epochs. The loss function combined the cross-entropy classification objective with optional physiological regularizers when enabled.

For comparative performance evaluation, five pretrained CNNs—-SqueezeNet, VGG-16, GoogLeNet, AlexNet, and ShuffleNet-—were employed as baseline models. The training parameters for all pretrained CNNs were set identical to those used for CardioPulmoNet to ensure fair comparison under consistent experimental conditions. Furthermore, linear SVMs were utilized for image classification using features extracted from the pre-trained CNNs and CardioPulmoNet. This hybrid approach enabled assessment of the discriminative power of learned representations across deep feature extractors and physiologically inspired architectures.

Model performance was evaluated using accuracy (ACC), sensitivity (SEN), specificity (SPE), precision (PRE), F1 score, Matthews correlation coefficient (MCC), Jaccard index (JI), and the area under the receiver operating characteristic curve (AUC). Each result was averaged across ten folds in a cross-validation scheme.

### 3.1 Oral Squamous Cell Carcinoma Dataset

Oral squamous cell carcinoma (OSCC) represents the most prevalent form of oral cancer, characterized by malignant epithelial proliferation and potential invasion into adjacent connective tissues. Accurate histopathological classification of OSCC is clinically critical, as it guides early diagnosis, treatment planning, and prognosis assessment.

The dataset used in this study comprises histopathological images of normal and cancerous oral tissues. The data were originally reported in [32]. It is publicly available from the Mendeley Data repository [33]. The images are organized into two subsets captured at different magnifications. One subset contains 89 images of normal oral epithelium and 439 images of oral OSCC tissue sections at 100*×* magnification. All images were acquired using a Leica ICC50 HD microscope from hematoxylin and eosin (H&E) stained slides, which were collected, prepared, and annotated by medical experts from 230 patients.

For the classification experiments presented in this work, a balanced subset comprising 89 images of normal oral epithelium and 89 images of OSCC was selected to ensure fair evaluation and mitigate class imbalance effects. Figure 2 illustrates representative H&E-stained histopathological images of normal oral epithelium and OSCC. Table 1 presents the performance metrics obtained for the pretrained CNNs, CardioPulmoNet, and their combinations with linear SVMs.

**Table 1:** Performance metrics obtained from different classifiers using OSCC data.

| Model | ACC | SEN | SPE | PRE | F1 | MCC | JI | AUC |
| --- | --- | --- | --- | --- | --- | --- | --- | --- |
| SqueezeNet | 0.76 | 0.74 | 0.78 | 0.77 | 0.75 | 0.52 | 0.61 | 0.87 |
| SqueezeNet-SVM | 0.79 | 0.80 | 0.78 | 0.78 | 0.79 | 0.57 | 0.65 | 0.90 |
| VGG-16 | 0.74 | 0.63 | 0.85 | 0.81 | 0.71 | 0.50 | 0.55 | 0.88 |
| VGG-16-SVM | 0.78 | 0.70 | 0.85 | 0.83 | 0.76 | 0.56 | 0.61 | 0.89 |
| GoogLeNet | 0.84 | 0.79 | 0.90 | 0.89 | 0.84 | 0.70 | 0.72 | 0.95 |
| GoogLeNet-SVM | 0.83 | 0.91 | 0.75 | 0.79 | 0.84 | 0.67 | 0.73 | 0.93 |
| AlexNet | 0.88 | 0.88 | 0.88 | 0.88 | 0.88 | 0.77 | 0.79 | 0.98 |
| AlexNet-SVM | 0.90 | 0.88 | 0.92 | 0.92 | 0.90 | 0.80 | 0.81 | 0.98 |
| ShuffleNet | 0.78 | 0.80 | 0.76 | 0.77 | 0.78 | 0.56 | 0.65 | 0.90 |
| ShuffleNet-SVM | 0.79 | 0.81 | 0.78 | 0.78 | 0.80 | 0.58 | 0.66 | 0.91 |
| CardioPulmoNet | 0.80 | 0.65 | 0.93 | 0.91 | 0.76 | 0.61 | 0.61 | 0.82 |
| CardioPulmoNet-SVM | 0.84 | 0.80 | 0.88 | 0.87 | 0.83 | 0.68 | 0.71 | 0.91 |

**Figure 2.**
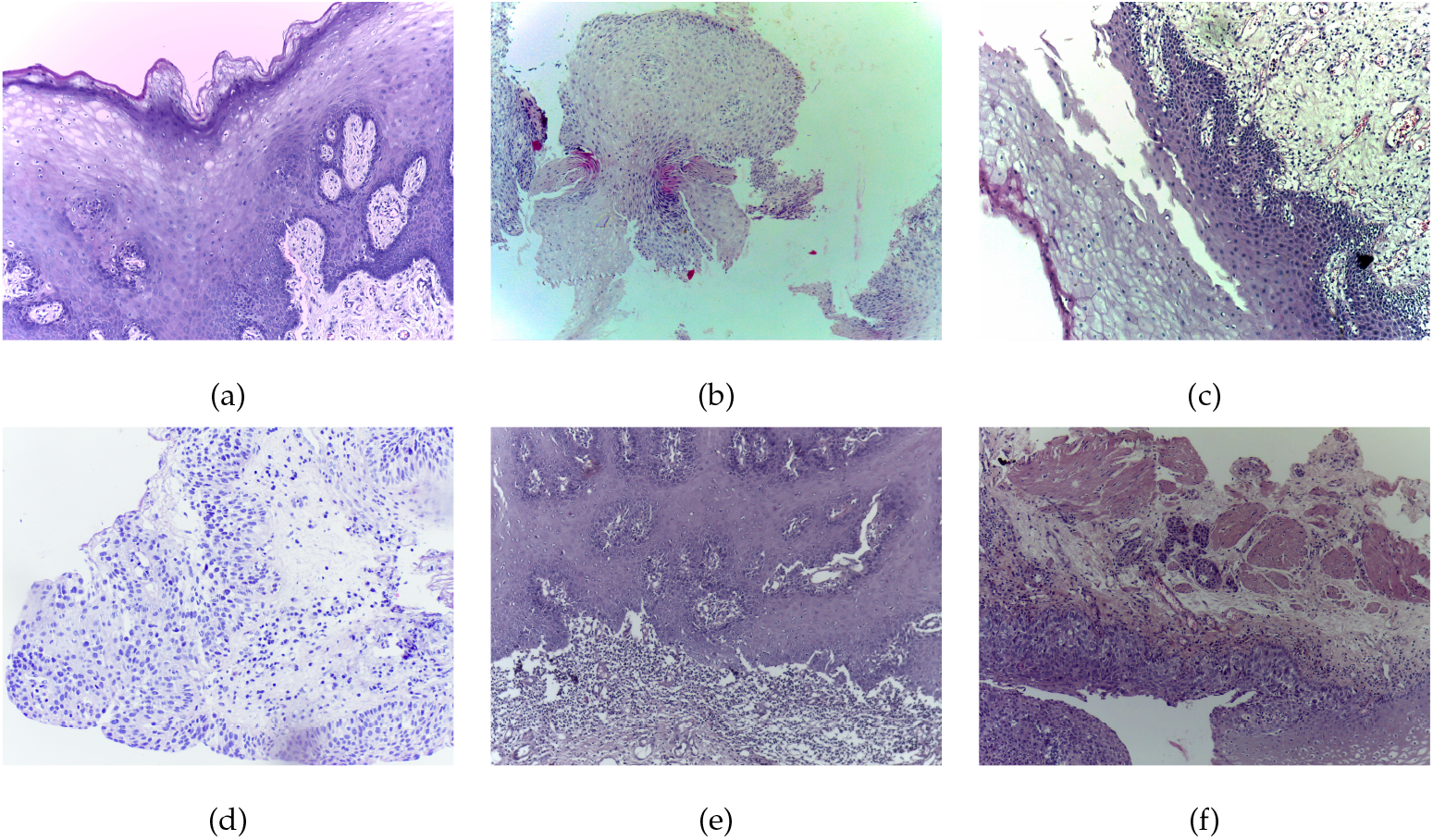
Representative H&E-stained histopathological images from the OSCC dataset: (a)-(c) normal oral epithelium, and (d)-(f) OSCC tissue.

### 3.2 Oral Submucous Fibrosis Dataset

Oral submucous fibrosis (OSMF) is a chronic, progressive, and potentially malignant disorder of the oral cavity characterized by epithelial atrophy, fibrosis of the submucosa, and restricted mouth opening. It is widely recognized as an oral precancerous condition with a measurable risk of transformation into OSCC. Accurate histopathological differentiation between normal and OSMF tissue is therefore essential for early detection, risk stratification, and timely clinical intervention. Automated image-based classification techniques provide a scalable and objective means of assisting pathologists, reducing inter-observer variability, and facilitating large-scale screening in resource-limited settings. Moreover, establishing robust models for OSMF classification serves as a critical step toward understanding disease progression and building predictive frameworks for malignant transformation in oral pathology.

The Oral Cancer Histology Image Database (ORCHID) [34] is a curated and publicly available resource designed to facilitate research in AI–based histopathological image analysis of oral cancer and precancerous lesions. The ORCHID database comprises a large, multicenter collection of high-resolution digitized slides captured at an effective magnification of 1000*×* (using a 100*×* objective lens). It includes histological images spanning multiple diagnostic categories, such as normal oral mucosa, OSMF, and OSCC.

All samples were H&E-stained and collected from patients under expert pathological supervision. The image acquisition process involved systematic digitization of tissue sections, producing non-overlapping image tiles at 1000*×* magnification to ensure uniform spatial resolution across samples. The complete dataset is publicly accessible through the Zenodo repository [35].

For the binary classification experiments presented in this study, 100 images of normal oral mucosa and 100 images of OSMF tissue were selected from the ORCHID dataset to evaluate the discriminative performance of the baseline and proposed models. Figure 3 illustrates representative H&E-stained histopathological images of normal oral mucosa and OSMF tissue. Table 2 summarizes the performance metrics obtained for the five pretrained CNNs, the proposed CardioPulmoNet model, and their combinations with linear SVMs.

**Table 2:** Performance metrics obtained from different classifiers using OSMF data.

| Model | ACC | SEN | SPE | PRE | F1 | MCC | JI | AUC |
| --- | --- | --- | --- | --- | --- | --- | --- | --- |
| SqueezeNet | 0.97 | 0.95 | 0.99 | 0.99 | 0.97 | 0.94 | 0.94 | 0.98 |
| SqueezeNet-SVM | 0.99 | 0.99 | 0.99 | 0.99 | 0.99 | 0.98 | 0.98 | 0.99 |
| VGG-16 | 0.89 | 0.95 | 0.82 | 0.84 | 0.89 | 0.78 | 0.81 | 0.95 |
| VGG-16-SVM | 0.94 | 1 | 0.88 | 0.89 | 0.94 | 0.89 | 0.94 | 0.98 |
| GoogLeNet | 0.99 | 0.99 | 0.99 | 0.99 | 0.99 | 0.98 | 0.98 | 0.99 |
| GoogLeNet-SVM | 0.99 | 0.98 | 0.99 | 0.99 | 0.98 | 0.97 | 0.98 | 0.99 |
| AlexNet | 0.99 | 0.98 | 1 | 1 | 0.99 | 0.98 | 0.98 | 0.99 |
| AlexNet-SVM | 0.99 | 1.0 | 0.99 | 0.99 | 0.99 | 0.99 | 0.99 | 1 |
| ShuffleNet | 0.99 | 0.99 | 0.99 | 0.99 | 0.99 | 0.98 | 0.98 | 0.99 |
| ShuffleNet-SVM | 0.99 | 0.98 | 1 | 1 | 0.99 | 0.98 | 0.98 | 1 |
| CardioPulmoNet | 0.70 | 0.70 | 0.70 | 0.70 | 0.70 | 0.40 | 0.54 | 0.89 |
| CardioPulmoNet-SVM | 1 | 1 | 1 | 1 | 1 | 0.99 | 0.99 | 1 |

**Figure 3.**
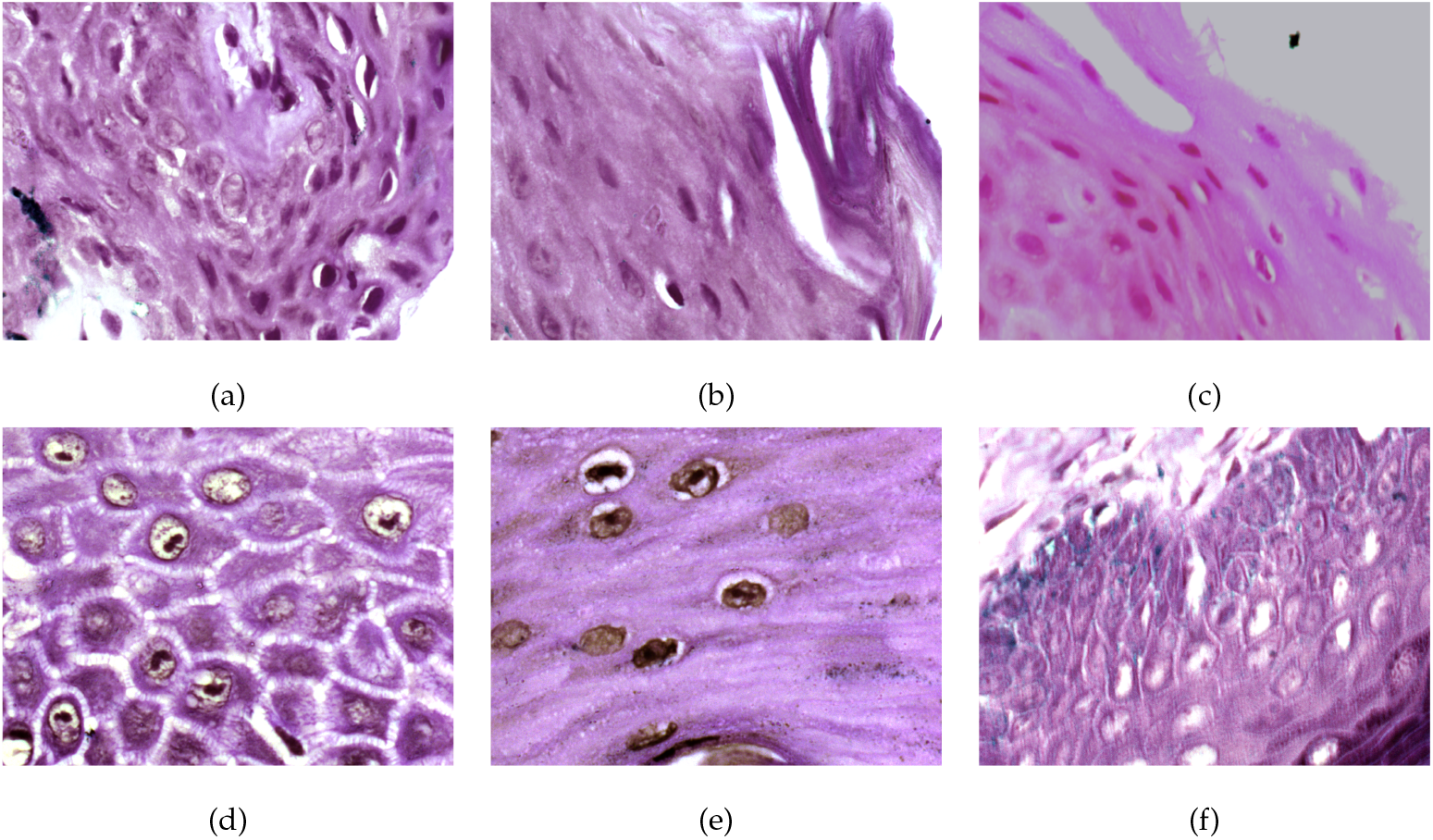
Representative H&E-stained histopathological images from the OSMF dataset: (a)-(c) normal oral mucosa, and (d)-(f) OSMF-affected tissue.

### 3.3 Heart Failure Dataset

The identification of patients with clinical heart failure from H&E-stained whole-slide histopathological images is of critical diagnostic and prognostic importance. Heart failure represents a complex clinical syndrome arising from diverse structural and functional myocardial abnormalities, and its progression is often accompanied by subtle histopathological changes, including myocyte hypertrophy, interstitial fibrosis, and inflammatory infiltration. Accurate recognition of these microscopic alterations is essential for confirming disease etiology, guiding therapeutic decisions, and assessing myocardial remodeling in both clinical and research settings. However, manual interpretation of whole-slide images is labor-intensive, subjective, and limited by inter-observer variability. Automated image-based classification can provide a standardized and objective approach for detecting histological signatures of heart failure, enabling scalable screening, improved diagnostic reproducibility, and potential integration into precision cardiopathology workflows.

The heart failure histopathology dataset [36] comprises H&E-stained sections of left ventricular myocardium obtained from both failing and non-failing human hearts. The failing cohort includes patients with clinically confirmed ischemic or idiopathic dilated cardiomyopathy who underwent heart transplantation or left ventricular assist device implantation. The non-failing cohort consists of organ donors with no documented history of cardiac dysfunction, whose hearts were not used for transplantation for non-pathological reasons.

All tissue samples were processed following standardized histological protocols, sectioned, stained with H&E, and digitized as whole-slide images at high resolution. Each whole-slide image was subsequently downsampled to an effective magnification of 5*×* to balance tissue coverage and computational efficiency. From these slides, eleven non-overlapping regions of interest, each measuring 2500 *µ*m^2^, were randomly selected to capture representative myocardial morphology. The resulting dataset provides a balanced and well-curated histopathological resource for studying structural alterations associated with clinical heart failure, including myocyte hypertrophy, fibrosis, and inflammatory infiltration. All image data are publicly accessible through the Image Data Resource under accession number `idr0042` (https://idr.openmicroscopy.org/).

In this study, a total of 200 images were selected for the classification task, comprising 100 images from each cohort. Figure 4 shows representative H&E-stained histopathological sections of non-failing and heart failure myocardium. Table 3 summarizes the performance metrics achieved by the pretrained CNNs, CardioPulmoNet, and their combinations with linear SVMs.

**Table 3:** Performance metrics obtained from different classifiers using heart failure data.

| Model | ACC | SEN | SPE | PRE | F1 | MCC | JI | AUC |
| --- | --- | --- | --- | --- | --- | --- | --- | --- |
| SqueezeNet | 0.86 | 0.91 | 0.81 | 0.83 | 0.87 | 0.72 | 0.76 | 0.95 |
| SqueezeNet-SVM | 0.86 | 0.79 | 0.93 | 0.92 | 0.85 | 0.73 | 0.74 | 0.97 |
| VGG-16 | 0.68 | 0.81 | 0.55 | 0.64 | 0.72 | 0.37 | 0.56 | 0.81 |
| VGG-16-SVM | 0.60 | 0.48 | 0.71 | 0.62 | 0.54 | 0.20 | 0.37 | 0.83 |
| GoogLeNet | 0.86 | 0.90 | 0.81 | 0.83 | 0.86 | 0.71 | 0.76 | 0.94 |
| GoogLeNet-SVM | 0.81 | 0.87 | 0.76 | 0.78 | 0.82 | 0.63 | 0.70 | 0.92 |
| AlexNet | 0.85 | 0.89 | 0.81 | 0.82 | 0.86 | 0.70 | 0.75 | 0.97 |
| AlexNet-SVM | 0.89 | 0.92 | 0.86 | 0.87 | 0.89 | 0.78 | 0.81 | 0.97 |
| ShuffleNet | 0.87 | 0.84 | 0.91 | 0.90 | 0.87 | 0.75 | 0.77 | 0.95 |
| ShuffleNet-SVM | 0.88 | 0.88 | 0.89 | 0.89 | 0.88 | 0.77 | 0.79 | 0.96 |
| CardioPulmoNet | 0.75 | 0.80 | 0.70 | 0.73 | 0.76 | 0.50 | 0.62 | 0.85 |
| CardioPulmoNet-SVM | 0.91 | 0.95 | 0.86 | 0.87 | 0.91 | 0.81 | 0.83 | 0.97 |

**Figure 4.**
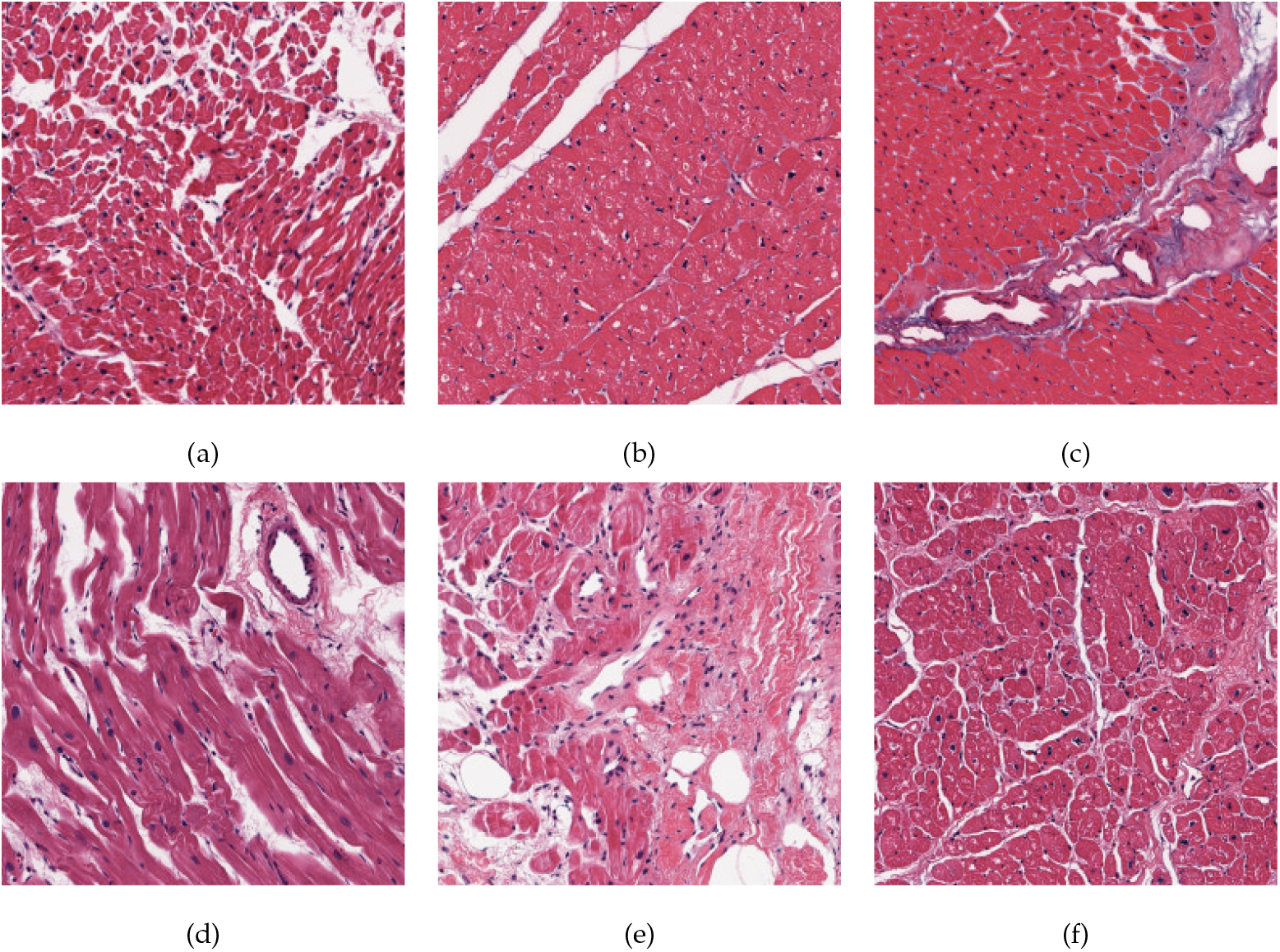
Representative H&E-stained histopathological images from the heart failure dataset: (a)–(c) normal cardiac tissue (non-failing hearts), and (d)–(f) cardiac tissue from patients with end-stage heart failure.

## 4 Discussion

The experimental results across three histopathological datasets (OSCC, OSMF, and heart failure) collectively demonstrate the effectiveness and generalizability of CardioPulmoNet as a physiologically grounded feature extractor. The model consistently produced competitive or superior performance compared to established pretrained CNNs, especially when its features were coupled with a linear SVM classifier.

For the classification of OSCC and normal oral tissue (Table 1), CardioPulmoNet achieved a strong baseline accuracy of 0.80 and specificity of 0.93 without pretraining, demonstrating its ability to learn discriminative and physiologically meaningful representations from limited data. When combined with an SVM, performance improved to an accuracy of 0.84 and an AUC of 0.91, indicating that the SVM enhanced the separability of latent features representing malignant versus normal epithelial morphology. Compared with pretrained CNNs such as AlexNet, GoogLeNet, and SqueezeNet, CardioPulmoNet-SVM achieved comparable or superior balanced performance while offering higher interpretability due to its biologically inspired dual-stream design.

For the OSMF dataset (Table 2), pretrained CNNs achieved near-perfect performance (ACC *>* 0.95, AUC ≈ 1.00), reflecting the high separability between normal and fibrotic tissues. Although CardioPulmoNet alone achieved moderate results (ACC = 0.70, AUC = 0.89), coupling its learned embeddings with an SVM led to perfect classification (ACC = 1.00, AUC = 1.00). This finding highlights that the model’s internal representations, while not convolutionally derived, are highly linearly separable, enabling complete discrimination between fibrotic and normal oral mucosa. The results further validate that CardioPulmoNet’s dual-stream architecture effectively captures the structural and textural cues underlying fibrotic pathology.

In the heart failure dataset (Table 3), pretrained CNNs achieved high accuracy (0.85– 0.89) and AUC values (0.94–0.97), reflecting their strong capability for general cardiac tissue characterization. CardioPulmoNet achieved a baseline accuracy of 0.75 and AUC of 0.85, but when paired with an SVM, performance improved markedly to an accuracy of 0.91, sensitivity of 0.95, and AUC of 0.97, matching or exceeding most pretrained CNN baselines. The improvement indicates that the model effectively encodes physiologically interpretable representations of myocardial remodeling and failure-related disorganization, while the linear classifier exploits the resulting separability for reliable prediction.

Taken together, these results confirm that CardioPulmoNet, even without pretraining, can generate biologically coherent, data-efficient, and generalizable feature representations across diverse histopathological domains. The combination with an SVM consistently enhances performance, suggesting that the model’s latent space is highly structured and discriminative. Importantly, CardioPulmoNet differs from conventional CNNs by embedding cardiopulmonary dynamics, ventilation and perfusion coupling, as an analogical learning framework that stabilizes feature exchange through homeostatic regularization. This biologically inspired mechanism contributes to the model’s robustness against small-sample variability, a critical limitation in medical imaging datasets.

Overall, the findings highlight a new paradigm for computational pathology, wherein biologically grounded architectures can achieve interpretability and high diagnostic accuracy without relying on large-scale pretraining. The strength of CardioPulmoNet lies in its explicit integration of physiological modeling with deep feature learning, offering multiple advantages that collectively support explainability, transferability, and clinical trustworthiness. First, by analogizing cardiopulmonary dynamics—the bidirectional exchange between pulmonary ventilation and cardiac perfusion—to dual feature streams, the network encodes interactions between local texture patterns and global structural context in a physiologically interpretable manner. This coupling enables the model to maintain equilibrium between high-level semantic representations, capturing diagnostically relevant tissue organization and pathological context, and fine-grained morphological details that characterize cellular and subcellular variations. By jointly preserving global semantic coherence and local textural precision, the model aligns its computational representations with biologically interpretable patterns observed in histopathology, thereby enhancing both diagnostic accuracy and interpretability.

Second, the incorporation of a homeostatic loss function imposes a biologically motivated equilibrium between feature subsystems, analogous to the physiological regulation of oxygen and blood flow. This regulatory mechanism enhances training stability and constrains the feature space to remain physiologically plausible, thereby improving both interpretability and robustness to data variability, a key challenge in histopathological imaging, where sample heterogeneity and limited data are common.

Third, because CardioPulmoNet operates without dependence on large-scale pretraining, it demonstrates strong generalization across distinct datasets (OSCC, OSMF, and heart failure), suggesting that its learned feature embeddings capture underlying morphological principles rather than dataset-specific artifacts. This data efficiency enhances transferability to new clinical contexts with minimal retraining.

Finally, the architecture’s modularity and physiologically interpretable design make its decision-making process more transparent to medical experts, facilitating integration into diagnostic workflows and regulatory frameworks that increasingly emphasize explainable AI. Together, these characteristics establish CardioPulmoNet as a promising foundation for next-generation computational pathology systems that are scientifically interpretable, clinically adaptable, and aligned with the principles of trustworthy AI in medicine.

## 5 Conclusion

This study introduced CardioPulmoNet, a physiologically grounded neural architecture designed to model cardiopulmonary dynamics for histopathological image classification. By conceptualizing pulmonary ventilation and cardiac perfusion as interacting feature streams, the network learns complementary local and global representations that promote balanced and interpretable feature learning. Across three datasets—oral squamous cell carcinoma, oral submucous fibrosis, and heart failure—CardioPulmoNet demonstrated competitive performance relative to established pretrained convolutional models, particularly when its learned features were classified using linear support vector machines.

The results suggest that embedding physiological analogies within neural architectures can enhance both interpretability and data efficiency, offering an alternative to conventional purely data-driven approaches. The combination of biological grounding and deep feature learning provides a principled framework for developing explainable and transferable AI systems in computational pathology.

Further investigation is required to develop the scalability of CardioPulmoNet on multimodal diagnostic data—including genomics, physiological signals, and clinical text. Future studies should also examine its potential as a pretrained foundation model capable of domain adaptation across diverse disease contexts. Such extensions will be essential for validating its translational utility and for establishing physiologically inspired deep learning as a reliable and trustworthy paradigm in medical AI.

## Data Availability

All 3 datasets produced are available online provided earlier in other section.

https://data.mendeley.com/datasets/ftmp4cvtmb/2

https://doi.org/10.5281/zenodo.12636426

https://idr.openmicroscopy.org/webclient/

